# Research topics and trends of lumbar spondylolisthesis: a bibliometric study with machine-learning based text mining

**DOI:** 10.1101/2022.05.25.22275576

**Authors:** Guoxin Fan, Jiaqi Qin, Yufeng Li, Sheng Yang, Longfei Huang, Huaqing Liu, Shisheng He, Xiang Liao

**Affiliations:** Department of Pain Medicine, Huazhong University of Science and Technology Union Shenzhen Hospital, Shenzhen, 518060, China; Guangdong Key Laboratory for Biomedical Measurements and Ultrasound Imaging, School of Biomedical Engineering, Shenzhen University Health Science Center, Shenzhen 518060, China; Artificial Intelligence Innovation Center, Research Institute of Tsinghua, Pearl River Delta, Guangzhou, 510735, China; Department of Orthopedics, The Eighth Affiliated Hospital Sun Yat-sen University, Shenzhen, 518033, China; Department of Orthopedics, Shanghai Tenth People’s Hospital, Tongji University School of Medicine, Shanghai, 200072, China; Spinal Pain Research Institute, Tongji University School of Medicine, Shanghai, 200072, China; Department of Orthopedics, Nanchang Hongdu Hospital of Traditional Chinese Medicine, 1399 Diezihu Road, Donghu District, Nanchang, 330006, China

**Keywords:** Bibliometrics, Lumbar Spondylolisthesis, Machine Learning, Latent Dirichlet allocation, Text Minging

## Abstract

**Objectives:** The study aimed to conduct a bibliometric analysis of publications concerning lumbar spondylolisthesis, as well as explore its research topics and trends with machine-learning based text mining.

**Methods:** The data were extracted from the Web of Science Core Collection (WoSCC) database and analyzed in Rstudio1.3.1. Annual publication production and the top 20 productive authors over time were presented. Additionally, top 20 productive journals and top 20 impact journals were compared by spine-subspecialty or not. Similarly, top 20 productive countries/regions and top 20 impact countries/regions were compared by developed countries/regions or not. The collaborative relationship among countries and the research trends in the past decade were presented by R package “Bibliometrix”. Latent Dirichlet allocation (LDA) analysis was conducted to classify main topics of lumbar spondylolisthesis.

**Result:** Up to 2021, a total number of 4990 articles concerning lumbar spondylolisthesis were finally included for analysis. Spine-subspecialty journals were found to be dominant in productivity and impact of the field, and SPINE, EUROPEAN SPINE JOURNAL and JOURNAL OF NEUROSURGERY-SPINE were the top 3 productive and the top 3 impact journals in this field. USA, China and Japan have contributed to over half of the publication productivity, but European countries seemed to publish more influential articles. It seemed that developed countries/regions tended to produce more articles as well as high influential articles, and international collaborations focused on USA, Europe and eastern Asia. Publications concerning emerging surgical technique was the major topic, followed by radiographic measurement and epidemiology for this field. Cortical bone trajectory, oblique lumbar interbody fusion, oblique lateral lumbar interbody fusion, lateral lumbar interbody fusion, degenerative lumbar spondylolisthesis, adjacent segment disease, spinal canal stenosis, minimally invasive transforaminal lumbar interbody fusion and percutaneous pedicle screw were the recent hotspots over the past 5 years.

**Conclusions:** The study successfully summarized the productivity and impact of different countries/regions and journals, which should benefit the journal selection and pursuit of international collaboration for researcher who were interested in the field of lumbar spondylolisthesis. Additionally, the current study may encourage more researchers in the field and somewhat inform their research direction in the future.

## Introduction

Lumbar spondylolisthesis, especially in the elderly, is one of the most common causes of low back pain(1). Lumbar spondylolisthesis is defined as a slip of one vertebra over the other beneath, causing instability of the segment and compression of the cauda equina(2). The incidence of lumbar spondylolisthesis is 4-6% in the general population(3), which typically occurs in the occurs at the fourth and fifth lumbar vertebrae(4). In the United States, approximately 11.5% of the population suffers from this disease(5). Based on its etiology, lumbar spondylolisthesis can be divided into isthmic or degenerative spondylolisthesis. The former could be post-traumatic fractures in the pars interarticularis or congenital defects, while the latter is a result of disorders of the disc space or degenerative arthritis(6). Currently, the risk factors for spondylolisthesis are considered to be older age, female gender, larger body mass index and sagittal facet orientation(7).

According to the current agreement, the treatment of lumbar spondylolisthesis essentially incorporates conservative treatment or surgical treatment. The nonsurgical treatment options may include physical therapy, exercise, epidural steroid injections for pain, and medications. If conservative therapy fails, a surgical intervention was usually recommended(8). Surgical options include either decompression alone or decompression with fixation and fusion, with the essential objectives of neural decompression and stability reconstructions as well as restoration of sagittal alignment(9). Recently, minimally invasive spine surgery has been widely adopted in surgical management of lumbar spondylolisthesis(10-12). Therefore, probing and summarizing the research topics or trends of lumbar spondylolisthesis may benefit potential doctors and researchers who were interested in this field.

Bibliometric analysis is a powerful tool to depict the research activities of a certain field in details(13). For example, bibliometrics analysis can quantify the contribution of a research field (including different countries, institutions, journals, or authors), and to identify the research trends or topics in a particular field(14). Nowadays, bibliometric analysis has been widely adopted in summarizing medical fields, like emerging techniques(15-17) and common disease(18-20). In spine field, bibliometric studies about cervical myelopathy(21), spinal stenosis(22), spinal cord injury(23), scoliosis(24) are thriving. However, no bibliometric study concerning lumbar spondylolisthesis was available. Meanwhile, latent Dirichlet allocation (LDA) is a popular machine learning algorithm that has been accepted as a bibliometric tool to obtain research topics for a specific field(25-27). Thus, the purpose of this work is to explore the research topics and trends of lumbar spondylolisthesis via bibliometric analysis.

## Methods

### Data Acquisition

Relevant literatures were collected from the Web of Science Core Collection (WoSCC) database. The search terms are “spondylolisthesis”. The time interval was set to 1975 to 2021. Only articles were included, and no language restrictions were applied. To avoid bias incurred by frequent database renewal, all literature retrieval and data downloads were completed in a single day, Jan 18, 2022. Considering that data were directly downloaded from the data set, ethical approval was not required. WoSCC data of full record and references (including titles, countries of origin, institutions, journals, authors, etc.) were extracted in bib format and then imported into the Rstudio1.3.1. for bibliometric analysis.

### Quantitative analysis

Annual publication production and the top 20 productive authors over time were presented. Additionally, top 20 productive journals and top 20 impact journals were compared by spine-subspecialty or not. Similarly, top 20 productive countries/regions and top 20 impact countries/regions were compared by developed countries/regions or not. The productivity of journals and countries/regions was measured by the total number of publications, whereas the impact of journals and countries/regions was assessed by H-index or average article citations (15). H-index was characterized as the extent that an entity has published at least h papers that have been each cited at least h times (28). All the data were analyzed by Rstudio1.3.1 and R package “Bibliometrix” was used to conduct quantitative analysis of different entity’s contributions.

### Research topics and trends

LDA can create a vocabulary of terms and then classify the included publications into different topics(18). We used the package “lda” in R language to conduct LDA analysis of included abstracts. The collaborative relationship among countries and the research trend in the past decade were presented by R package “Bibliometrix”.

## Results

### General information

Up to 2021, a total number of 4990 articles concerning lumbar spondylolisthesis were finally included for analysis. Average citations per documents reached 24.43, and average citations per year per documents reached 1.829. A total of 623 sources were identified while most sources were academic journals, as only a few articles were book chapters. Annual published articles concerning lumbar spondylolisthesis kept rising over time (**Figure 1**), and the publications of the latest 10 years contributed to 56.9% (2841/4990) productivity of this field. Similarly, almost all the top 20 productive authors published most of their articles concerning lumbar spondylolisthesis in years between 2010 and 2020.

**Figure 1.**
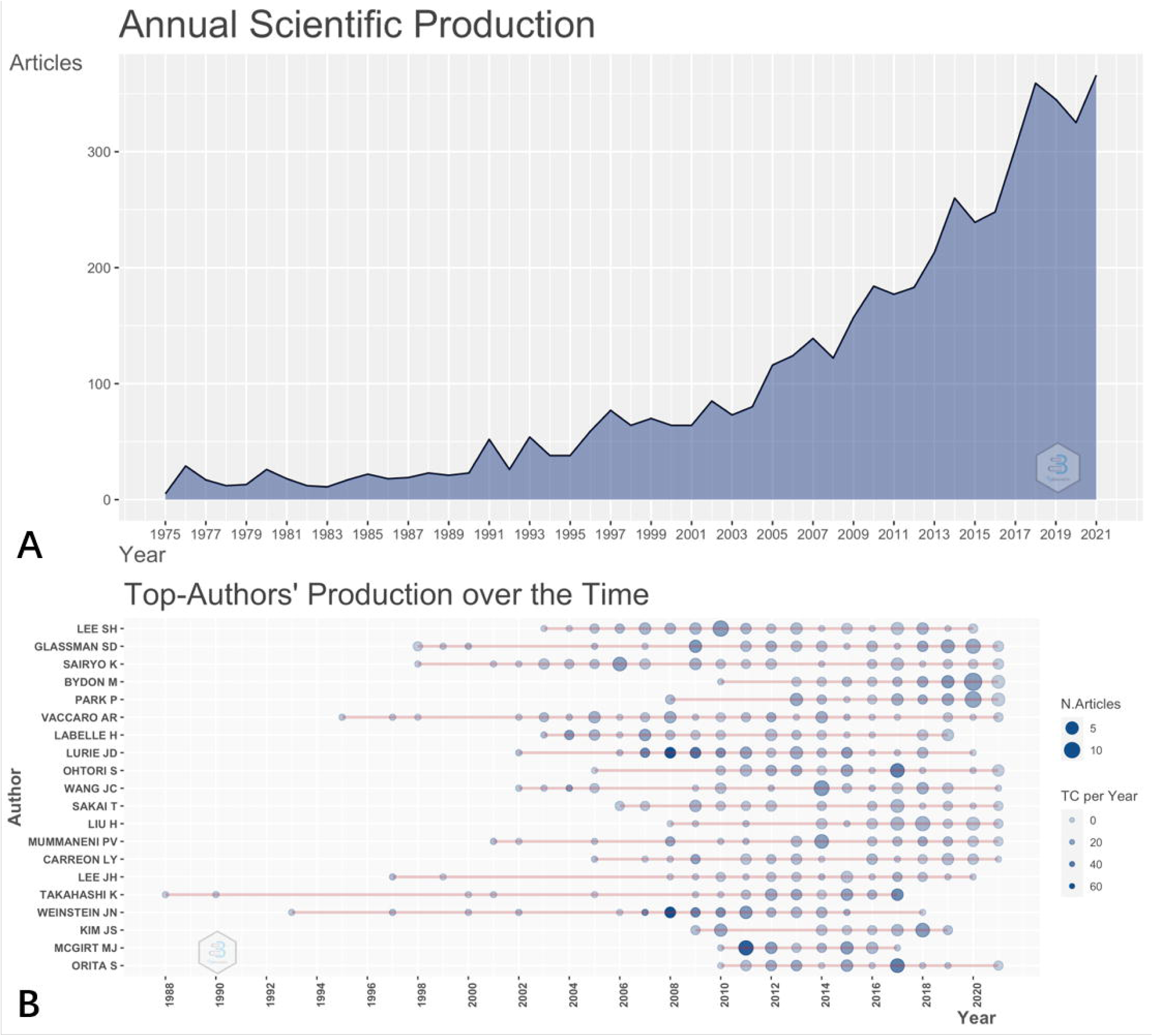
Annual publication production and the top 20 authors over time.

### Quantitative analysis

The top 20 productive journals and the top 20 impact journals publishing articles concerning lumbar spondylolisthesis were listed in **Table 1**. Articles in the top 20 journals (2789) were equal to 55.89% of all 4990 article publications. “SPINE” was the most productive journal (750 articles) and the most influential journal (H-index: 102) in the field. Similarly, “EUROPEAN SPINE JOURNAL” was the second productive (361 articles) and the second influential journal (H-index: 49), while “JOURNAL OF NEUROSURGERY-SPINE” was the third productive (255 articles) and the third influential journal (H-index: 47). Most articles (1952/2789) were published by spine-subspecialty journals among the top 20 productive journals, even though only 9 spine-subspecialty journals were in the top 20 productive lists. The top 3 influential journals all belonged to spine-subspecialty journals, although only one third of the top 20 influential journals were spine-subspecialty journals (7/20).

**Table 1.**
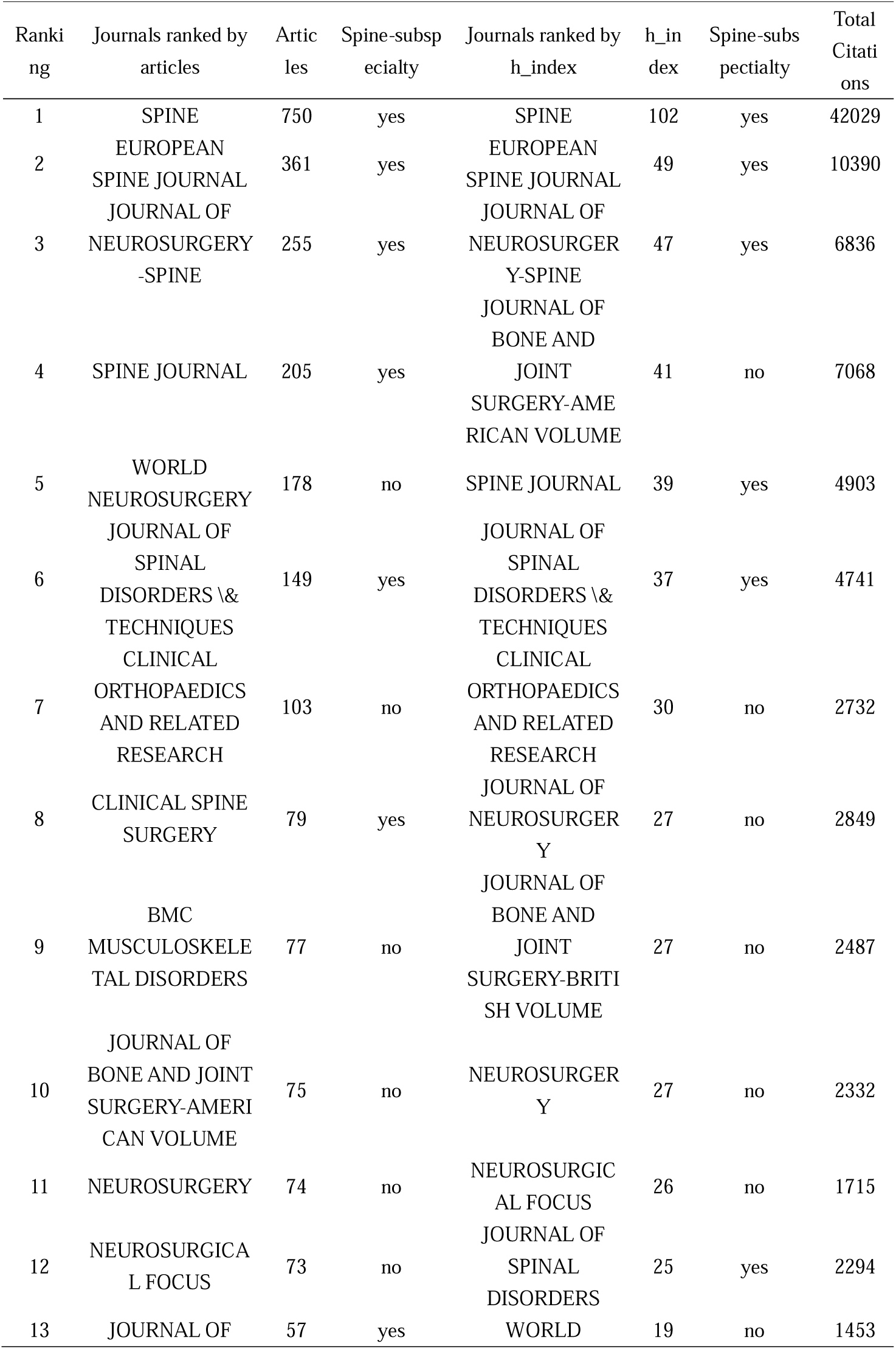
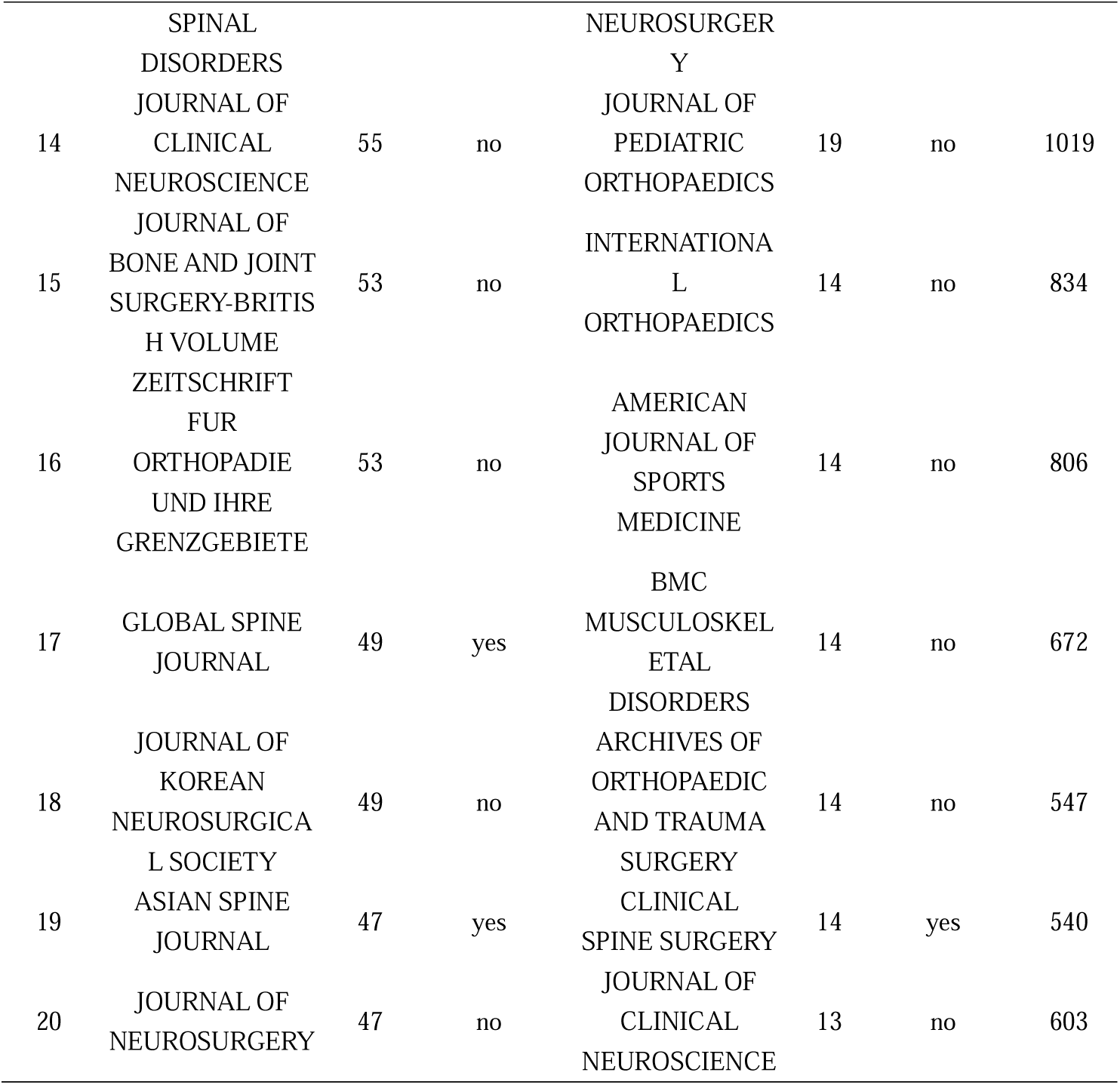
Top 20 productive and top 20 impact journals in the field of lumbar spondylolisthesis.

The top 20 productive and top 20 impact countries/regions publishing articles concerning lumbar spondylolisthesis were listed in **Table 2**. Articles in the top 20 countries/regions (4468) were equal to 89.54% of all 4990 article publications. USA was the most productive country (1570 articles) in this field, followed by China (536 articles) and Japan (531 articles). It was noted that Taiwan, as one of the most developed regions in China, accounted for nearly 20% of China’s productive contributions. However, only three non-developed countries were in the top 20 productive list, while developed countries/regions published 86.01% (3843/4468) articles among the top 20 productive lists. As we ranked the top impact countries/regions by average article citations, USA only ranked top 7 and Japan ranked top 16, while China was even not in the top 20 influential list. Among this list, only one country belonged to non-developed economic entity.

**Table 2.**
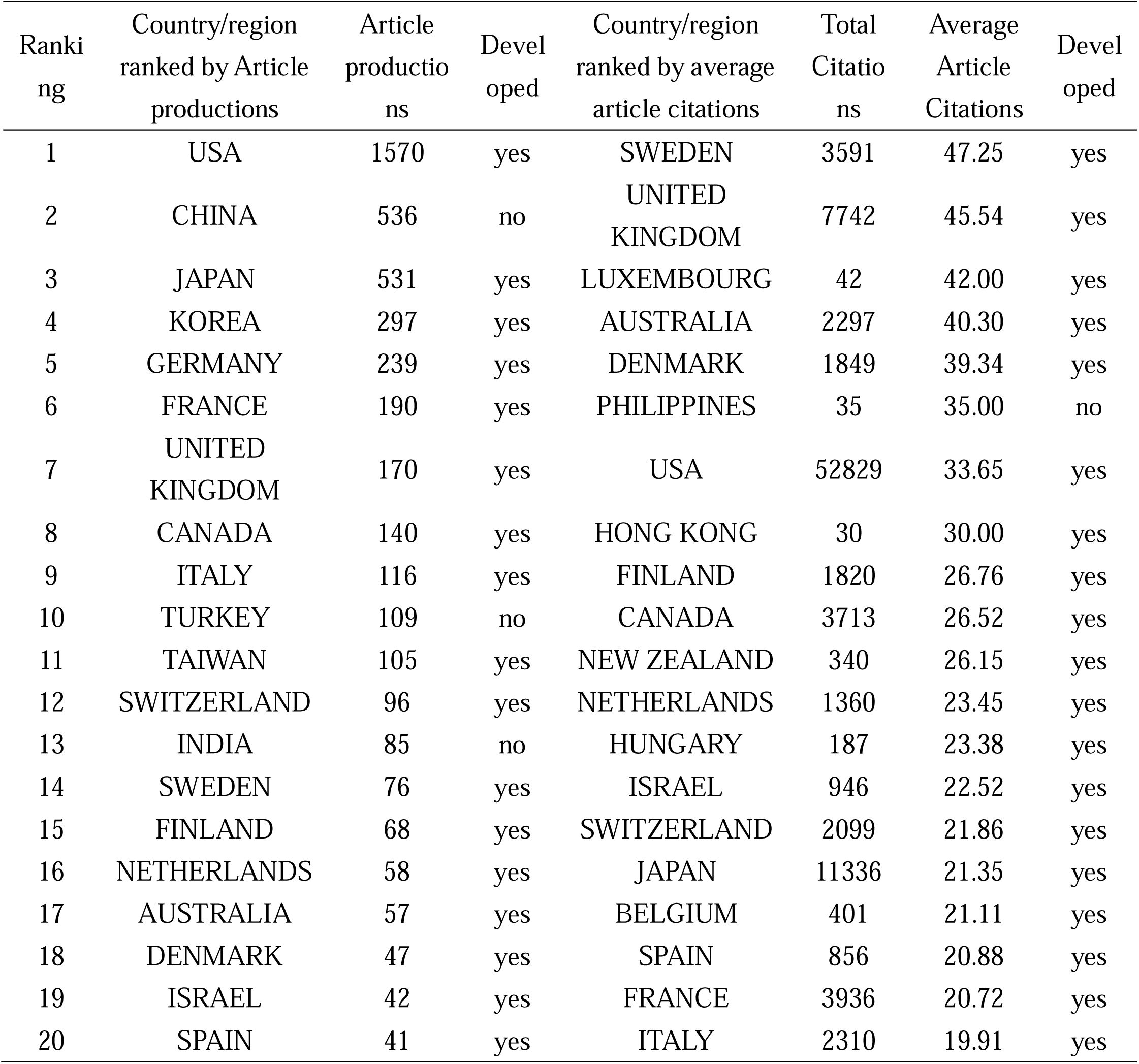
Top 20 productive and impact countries/regions in the field of lumbar spondylolisthesis.

| Ranking | Country/region ranked by Article productions | Article productions | Developed | Country/region ranked by average article citations | Total Citations | Average Article Citations | Developed |
| --- | --- | --- | --- | --- | --- | --- | --- |
| 1 | USA | 1570 | yes | SWEDEN | 3591 | 47.25 | yes |
| 2 | CHINA | 536 | no | UNITED KINGDOM | 7742 | 45.54 | yes |
| 3 | JAPAN | 531 | yes | LUXEMBOURG | 42 | 42.00 | yes |
| 4 | KOREA | 297 | yes | AUSTRALIA | 2297 | 40.30 | yes |
| 5 | GERMANY | 239 | yes | DENMARK | 1849 | 39.34 | yes |
| 6 | FRANCE | 190 | yes | PHILIPPINES | 35 | 35.00 | no |
| 7 | UNITED KINGDOM | 170 | yes | USA | 52829 | 33.65 | yes |
| 8 | CANADA | 140 | yes | HONG KONG | 30 | 30.00 | yes |
| 9 | ITALY | 116 | yes | FINLAND | 1820 | 26.76 | yes |
| 10 | TURKEY | 109 | no | CANADA | 3713 | 26.52 | yes |
| 11 | TAIWAN | 105 | yes | NEW ZEALAND | 340 | 26.15 | yes |
| 12 | SWITZERLAND | 96 | yes | NETHERLANDS | 1360 | 23.45 | yes |
| 13 | INDIA | 85 | no | HUNGARY | 187 | 23.38 | yes |
| 14 | SWEDEN | 76 | yes | ISRAEL | 946 | 22.52 | yes |
| 15 | FINLAND | 68 | yes | SWITZERLAND | 2099 | 21.86 | yes |
| 16 | NETHERLANDS | 58 | yes | JAPAN | 11336 | 21.35 | yes |
| 17 | AUSTRALIA | 57 | yes | BELGIUM | 401 | 21.11 | yes |
| 18 | DENMARK | 47 | yes | SPAIN | 856 | 20.88 | yes |
| 19 | ISRAEL | 42 | yes | FRANCE | 3936 | 20.72 | yes |
| 20 | SPAIN | 41 | yes | ITALY | 2310 | 19.91 | yes |

Collaborations among countries/regions and institutions in lumbar spondylolisthesis field were presented in **Figure 2**. Countries or region connected with red lines indicated there were some collaborations among them and the thickness of the red line was proportional to the number of collaborations. It seemed that USA was the hub country of publications, because there are lots of red lines radiated from USA to European countries, China, Korea, Japan and even Australia. Although India, Brazil, China, Japan and some middle eastern countries were also productive, there were little collaborations among them. As for institution collaboration, University of California-San Francisco was the most active institution in this field. It seemed that most active institutions in collaborations were from USA, followed by South Korea, and Canada. Although China was one of the most productive country, only two intuitions from Taiwan Province were recognized as the most active institutions in collaborations.

**Figure 2.**
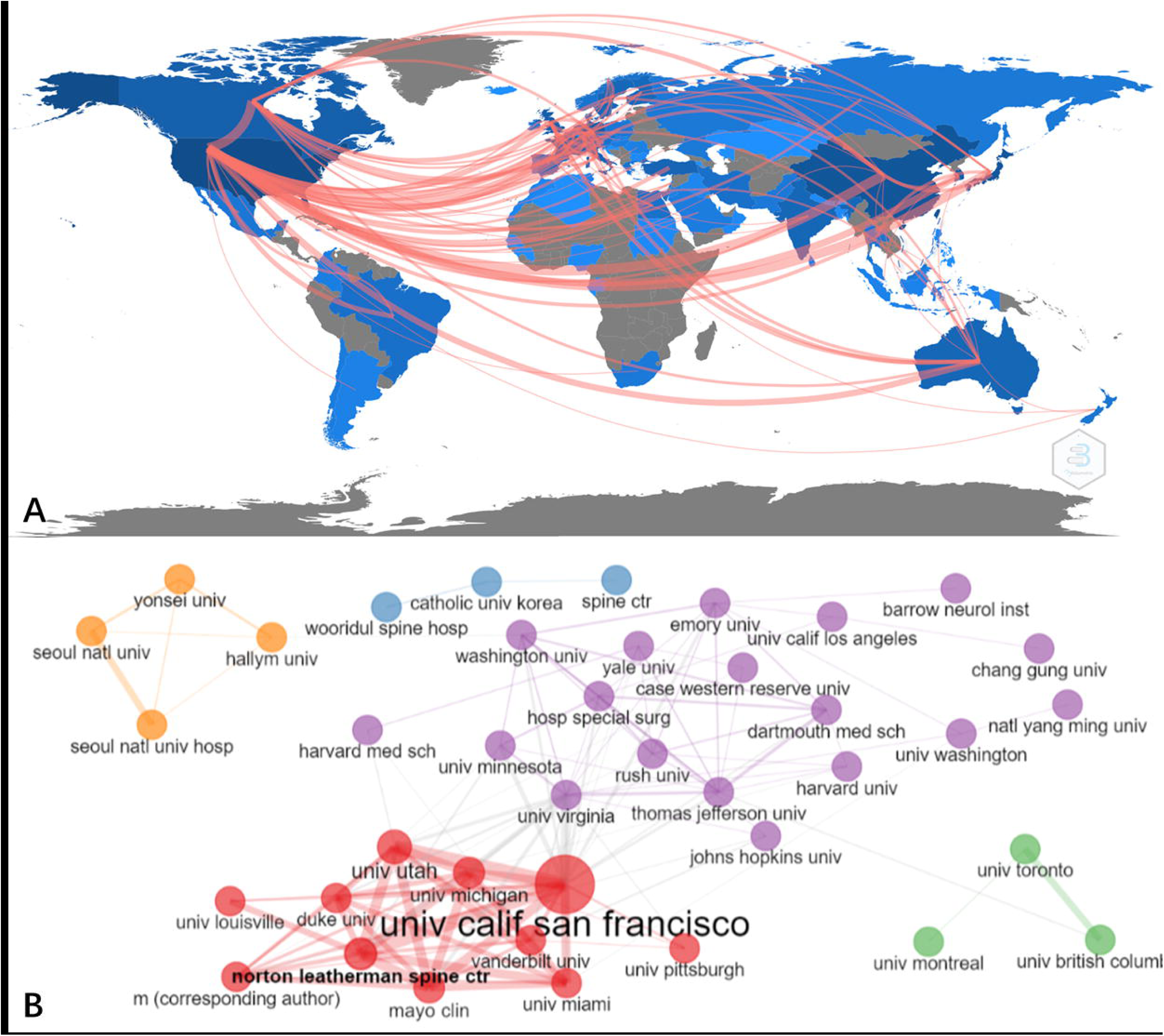
Collaborations in lumbar spondylolisthesis field. A: collaborations among countries/regions; B: collaborations among institutions.

## Research topics and trends

After removing articles without abstracts, a total of 4581 articles were finally included for LDA analysis. We found three major topics in this field, and named them as “Topic 1: surgery”, “Topic 2: radiology” and “Topic 3: epidemiology” (**Figure 3**). The publication proportion of these three topics were 37.2%, 34.3% and 28.5%, respectively. Additionally, we presented how the productivity of these three topics evolved over time. It seemed that one third of publications were concerned about the radiographic measurement of lumbar spondylolisthesis (Topic 2: radiology), and it kept as the most productive topic for two decades (1990 to 2010). After 2010, publications concerning all kinds of surgical management (Topic 1: surgery) became the dominant topic, and publications concerning prevalence, risk factors, quality of life and conservative treatment of lumbar spondylolisthesis (Topic 3: epidemiology) joined as another mainstream topic after 2015.

**Figure 3.**
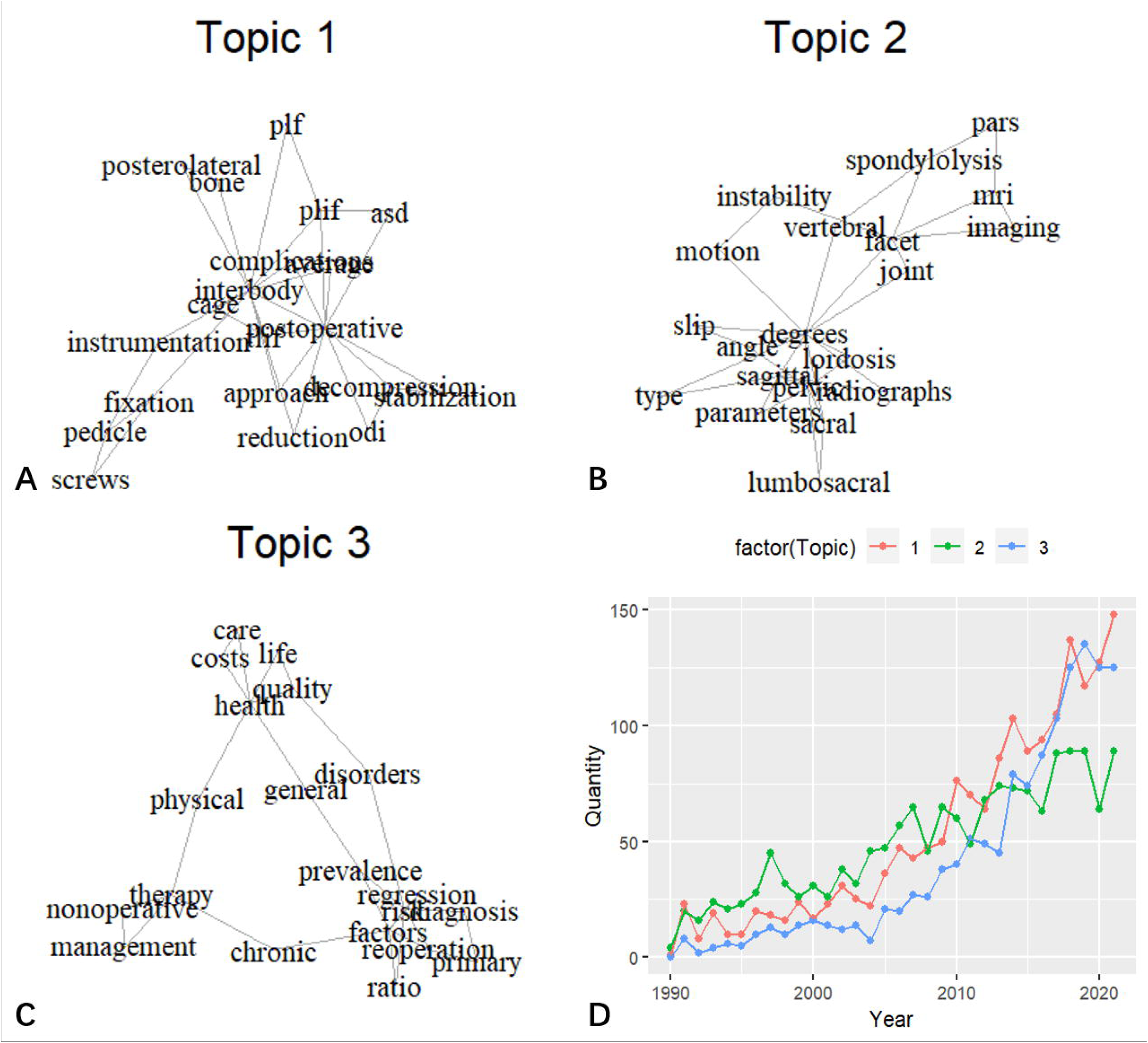
Research topics of lumbar spondylolisthesis over the past 3 decades. A: topic 1- surgery; B: topic 2- radiology; C:Topic 3: epidemiology; D: publication productivity of the three topics over the past three decades.

Using term frequency, we obtained the research trends of lumbar spondylolisthesis over the past one decade (**Figure 4**). Cortical bone trajectory (CBT), oblique lumbar interbody fusion (OLIF), oblique lateral lumbar interbody fusion (OLLIF), lateral lumbar interbody fusion (LLIF), degenerative lumbar spondylolisthesis, adjacent segment disease (ASD), spinal canal stenosis, minimally invasive transforaminal lumbar interbody fusion (MIS-TLIF) and percutaneous pedicle screw were the recent hotspots over the past 5 years.

**Figure 4.**
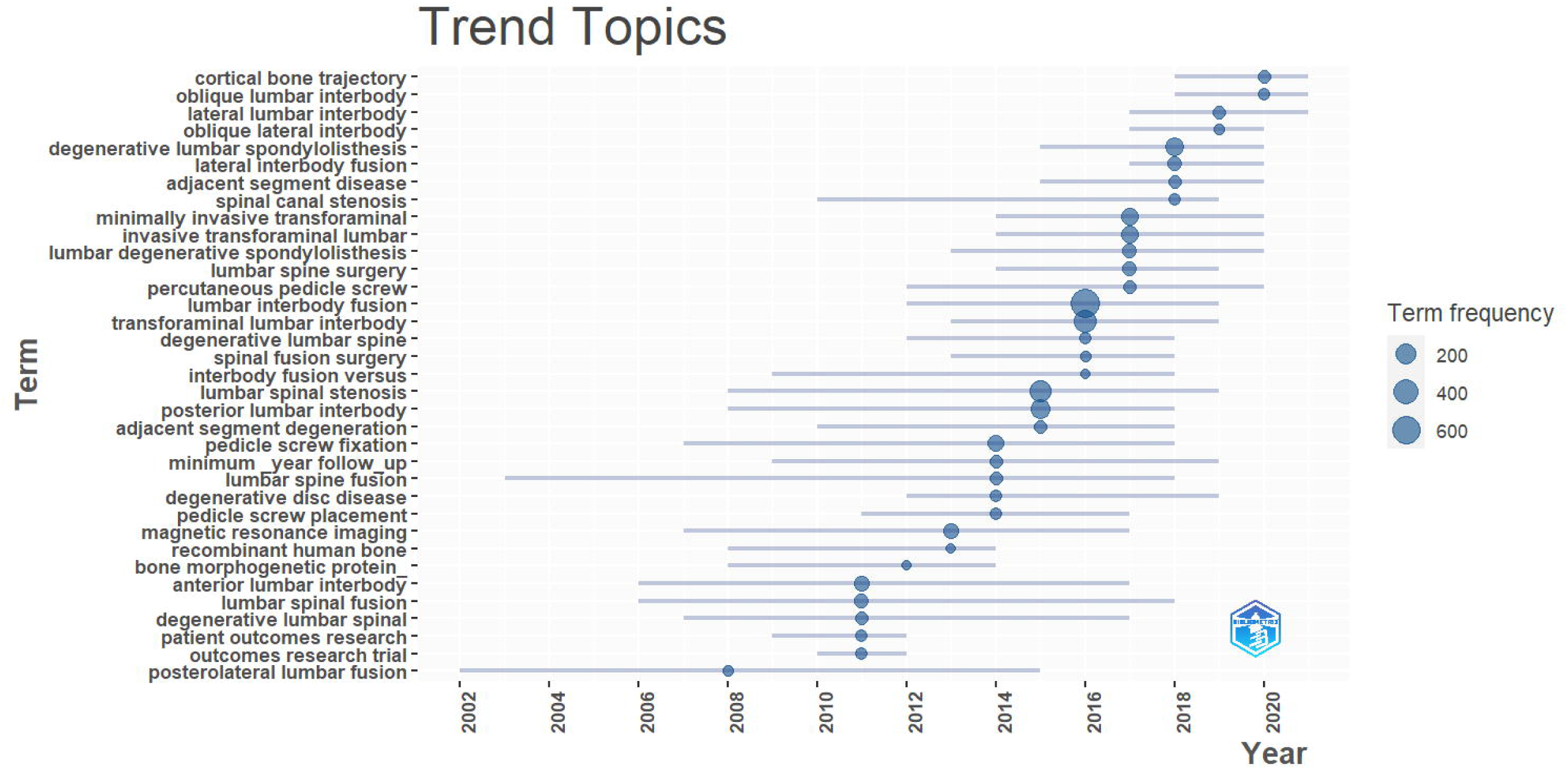
Research trends of lumbar spondylolisthesis.

## Discussion

Text mining and bibliometric analysis have been adopted widely in depicting a certain academic field in details. The current study quantified different entities’ academic contributions to lumbar spondylolisthesis, as well as provided an in-depth and visualized analysis of its topics and trends. To the best of our knowledge, this was the first bibliometric analysis of lumbar spondylolisthesis, which might help researchers gain a basic understanding, develop areas of focus and pursue further practice in this field.

Academic contributions of one field could be quantified by publication productivity and citation activity. The current study mainly quantified the contributions of different journals and countries in the field of lumbar spondylolisthesis. It seemed that SPINE, EUROPEAN SPINE JOURNAL and JOURNAL OF NEUROSURGERY-SPINE were the prior journals for researchers to follow or submit their own works, as these three journals were the top 3 productive and the top 3 impact journals in this field. Additionally, other spine-subspecialty journals like SPINE JOURNAL could be the second-line options, as spine-subspecialty journals were found to be dominant in productivity and impact of the field. For country/region contributions, USA, China and Japan have contributed to over half of the publication productivity. However, European countries seemed to publish more influential articles, as their average article citations were much higher. It seemed that developed countries/regions tended to produce more articles as well as high influential articles, as there were only a few non-developed countries/regions among the top 20 productive and the top 20 influential lists. Additionally, researchers may need to focus on USA, Europe and eastern Asia, if they need collaborations in the field of lumbar spondylolisthesis.

Natural language processing (NLP) is a popular research field where human language could be decoded by machine learning, which has been adopted to analyze academic publications recently(29). LDA is one of the most widely used machine learning algorithms in NLP(18, 25, 26), as it is scalable, computationally fast and close to what the human mind assigns while decoding text words. In the study, publications of Topic 2 remained as the dominant topic from 1990 to 2010, when all kinds of radiographic equipment spread over the world gradually. We assumed that the widespread of radiographic equipment enabled researchers to pay more attention to lumbar instability and thus identify key parameters of sagittal balance. These publications (Topic 2) did improve the knowledge of lumbar spondylolisthesis like classification(30) or sagittal balance(31), which was of great significance of guiding the management of lumbar spondylolisthesis(32-34). Publications of Topic 1 kept as the most productive topic after 2010, which could be explained by the emerging minimally invasive surgical technique and the lack of agreement on the best surgical approaches(35), as well as the controversy of the addition of fusion to decompression(36). Publications of Topic 3 gradually became the mainstream topic after 2015, which indicated the importance of systematic management of lumbar spondylolisthesis. As most spondylolisthesis patients were asymptomatic and only a few patients seeking treatment will have surgery, doctors from different subspecialties might hold different viewpoints about best options of nonoperative treatments, as well as their dosage and progression of physical therapy procedures(6). Failed conservative treatment would lead to more than double medical costs than those in the successfully treated cohort(37).

While major topics with historical perspective quickly informed potential researchers about the macroscopic picture of one academic field and how it evolved, recent hotspots might guide the future researches. The current study identified CBT, OLIF, OLLIF, LLIF, MIS-TLIF, percutaneous pedicle screw, degenerative lumbar spondylolisthesis, spinal canal stenosis and ASD were the research hotspots over the past 5 years. Interestingly, most of these hotspots (the first six) belong to minimally invasive surgical techniques, and all of them were derivatives of internal fixation or fusion technique instead of decompression. It was not a surprise as decompression technique has developed for decades, but internal fixation and fusion technique could be easily modified with different approaches and novel devices. Unlike the novel concept of the first six hotspots, spine physicians or surgeons might not be unfamiliar with degenerative lumbar spondylolisthesis, spinal canal stenosis and ASD. Degenerative lumbar spondylolisthesis became a hotspot instead of isthmic spondylolisthesis might be explained by growing aging population and higher prevalence of lumbar degeneration. Lumbar spondylolisthesis often accompanied by spinal canal stenosis and ASD is a common complication of spinal fusion, which indicated continuous attentions have been paid to these traditional problems.

Since most hotspots belonged to publications of Topic 1 (surgery), it was necessary to depict these surgical hotspots in details. CBT, as a caudal-to-rostral and medial-to-lateral pedicle screw insertion method, has merits of great fixation and pull-out strength even in osteoporosis patients due to its trajectory goes through cortical bone(38). Thus, CBT became popular in all kinds of spinal fusion including lumbar spondylolisthesis(39-41). Percutaneous pedicle screw is another widespread fixation technique with traditional trajectory, but its merits of minimally invasiveness popularized its utility in OLIF, OLLIF, LLIF and MIS-TLIF etc. OLIF takes an anterior retroperitoneal approach surgery through a small incision(42). Within the retroperitoneal space, surgeons will drag the psoas muscle backward, expose the intervertebral space between the psoas muscle and the abdominal aorta, and then perform decompression plus fusion, which is usually followed by posterior fixation of percutaneous pedicle screw. OLLIF is another minimally invasive fusion surgery described by AbbasiH in details in 2015(43). OLLIF uses the classic YESS endoscopy trajectory technology to establish a working channel, and then enters the intervertebral disc through Kambin’s triangle to complete discectomy and intervertebral fusion under full-endoscopy(44). LLIF or extreme lateral interbody fusion (XLIF) is another minimally invasive surgery by bluntly separate the psoas muscle to the lateral side of the intervertebral space(45, 46). It should be noted that muscle separation may damage the lumbar plexus, so intraoperative neuromonitoring is required for LLIF/XLIF. MIS-TLIF takes a small paramedial incision, and the ipsilateral articular process will be excised to expose the intervertebral foraminal window, through which decompression and fusion are performed(47). It is reported that MIS-TLIF has become one of the most widespread fusion surgeries(48).

The current study may have some limitations. First, we only analyzed articles from the WoSCC database, so publications not indexed in the WoSCC database were not considered and citation counts might be underestimated. Second, the study mainly analyzed some useful information (e.g. abstracts, titles, etc.) of the included publications instead of reviewing full texts. Last but may not least, as the WoSCC database kept updating and records of 2022 were not complete, we only analyzed the data by 2021, which might not reflect the most recent trends of 2022.

## Conclusions

The study successfully summarized the productivity and impact of different countries/regions and journals, which should benefit the journal selection and pursuit of international collaboration for researcher who were interested in the field of lumbar spondylolisthesis. Publications concerning emerging surgical technique was the major topic, followed by radiographic measurement and epidemiology for this field. CBT, OLIF, OLLIF, LLIF, MIS-TLIF, percutaneous pedicle screw, degenerative lumbar spondylolisthesis, spinal canal stenosis and ASD were the research hotspots of lumbar spondylolisthesis over the past 5 years. With macroscopic plus detailed analysis of publications concerning lumbar spondylolisthesis, the current study may encourage more researchers in the field and somewhat inform their research direction in the future.

## Data Availability

All data generated or analysed during this study are either included in this published article or its supplementary information fles.

## Declarations

### Ethics approval and consent to participate

Not applicable.

## Authors’ contributions

G.F. contributed to writing and data interpreting; J.Q. contributed to data analysis; Y.L. and S.Y. contributed to drafting and data extraction; H.L. and L.H. contributed to data analysis and critical revisions; X.L. and S.H. contributed to study design and administration support.

## Acknowledgements

Not applicable.

## Consent for publication

Not applicable.

